# COVID-19 Workplace Outbreaks by Industry Sector and their Associated Household Transmission, Ontario, Canada, January – June, 2020

**DOI:** 10.1101/2020.11.25.20239038

**Authors:** Michelle Murti, Camille Achonu, Brendan T. Smith, Kevin A. Brown, Jin Hee Kim, James Johnson, Saranyah Ravindran, Sarah A. Buchan

**Author notes:** **Corresponding Author:** Michelle Murti, 661 University Ave, Suite 1701, Toronto, ON, Canada, M5G1M1.

## Abstract

**Introduction:** Workplaces requiring in-person attendance of employees for ongoing operations may be susceptible to SARS-CoV-2 outbreaks that impact workers as well as their close contacts. To understand industry sectors impacted by workplace outbreaks in the first wave of the pandemic, and the additional burden of illness through household transmission, we analyzed public health declared workplace outbreaks between January 21 to June 30, 2020, and their associated cases from January 21 to July 28.

**Methods:** Number, size and duration of outbreaks were described by sector, and outbreak cases were compared to sporadic cases in the same time frame. Address matching identified household cases with onset ≥2 days before, ≥2 days after, or within 1 day of the workplace outbreak case.

**Results:** There were 199 outbreaks with 1245 cases, and 68% of outbreaks and 80% of cases belonged to i) Manufacturing, ii) Agriculture, Forestry, Fishing, Hunting, iii) Transportation and Warehousing. Median size of outbreaks was 3 cases (range: 1-140), and lasted median 7days (range: 0-119). Outbreak cases were significantly more likely to be male, younger, healthier, and have better outcomes. There were 608 household cases associated with 339 (31%) outbreak cases with valid addresses, increasing the burden of illness by 56%. The majority of household cases (368, 60%) occurred after the outbreak case.

**Conclusions:** Workplace outbreaks primarily occurred in three sectors. COVID-19 prevention measures should target industry sectors at risk by preventing introduction from exposed employees, spread in the workplace, and spread outside of the workplace.

**What is already known about this topic?:** COVID-19 outbreaks occur within workplaces and can spread to the community

**What is added by this report?:** From January 21 – June 30, 2020, there were 199 workplace outbreaks in Ontario, Canada; 68% of outbreaks and 80% of outbreak-associated COVID-19 case were in three industry sectors: Manufacturing, Agriculture/Forestry/Fishing/Hunting, and Transportation/Warehousing. Household transmission occurred among 31% of outbreak cases, resulting in a 56% increase in workplace outbreak-associated cases when burden of household transmission is considered.

**What are the implications for public health practice?:** Workplace outbreak prevention measures should be targeted to industry sectors at risk by preventing introduction from exposed employees, spread in the workplace, and transmission to the greater community.

## Introduction

The Coronavirus disease (COVID-19) pandemic caused by Severe Acute Respiratory Syndrome Coronavirus-2 (SARS-CoV-2) led to a lockdown on non-essential services in mid-March 2020 in Ontario, with subsequent staged regional re-opening of the economy from May 2020.^1,2^ Over this period, a number of workplace settings have been identified in Canada and internationally as particularly susceptible to outbreaks with high attack rates, such as food processing facilities.^3-7^ These outbreaks have highlighted the need for workplace infection and prevention and control measures to reduce the risk from workplaces, in particular where employees have close, prolonged contact.^8^ However, with re-opening of the economy and previously closed (“non-essential”) workplaces, ongoing assessment of outbreak settings is necessary to understand evolving COVID-19 transmission risk in workplaces. While other jurisdictions have identified higher risk industry sectors associated with outbreaks, similar systematic assessment is lacking in Canada.^5^

Assessment of the morbidity and mortality of cases associated with workplace outbreaks alone may under-represent the burden of illness associated with transmission from these outbreaks. Previous studies have shown the impacts from workplace outbreaks beyond the affected cases to their surrounding community adding to the overall burden of illness from household and community transmission.^3,9^ Identifying vulnerable industry sectors and their related community transmission is necessary to target public health interventions to prevent outbreaks and subsequent community spread. Our objectives were to: 1) describe workplace outbreaks in Ontario over the first six months of 2020 (the first wave of the SARS-CoV-2 pandemic and initial stages of re-opening); 2) describe cases associated with these workplace outbreaks; and 3) estimate the additional burden of illness due to associated household transmission.

## Methods

### Data Source

We obtained data on workplace outbreaks and laboratory-confirmed cases identified by the 34 local public health units (PHU) in Ontario.^10^ Data were obtained from the integrated Public Health Information System, the Toronto Public Health Coronavirus Rapid Entry System, the Ottawa Public Health COVID-19 Ottawa Database, the Middlesex-London COVID-19 Case and Contact Management tool and Ontario Case and Contact Management database, collectively known as CCM Plus. Cases were extracted from January 21, 2020, when COVID-19 became reportable in the province, to July 28, 2020, to account for the delay in surveillance and reporting of cases associated with workplace outbreaks declared January 21 - June 30, 2020.

### Outbreak definitions

PHUs are responsible for declaring outbreaks of SARS-CoV-2 in various settings, including workplaces, based on their assessment of risk of acquisition and transmission.^11^ Provincial guidance on the definition of a workplace outbreak (two or more cases reasonably acquired in the workplace) was only issued on June 11, 2020, and outbreaks with a single case were declared prior to this guidance based on health unit assessment of risk of an outbreak in the setting.^12^ In this report, workplaces refer to non-hospital, non-congregate living, and non-childcare settings. Agriculture outbreaks were excluded as congregate living settings if >4 cases had the same address. Workplace outbreak locations (address and business name as entered by the PHU) were manually classified according to the North American Industry Classification System (NAICS) into one of 20 industry sectors.^13^ Outbreak size is the number of confirmed cases linked by the PHU to an outbreak. Outbreak duration is the time from the accurate episode date of the first to the last case in the outbreak, and zero in outbreaks with a single case. Accurate episode date is determined by a hierarchy of symptom onset, specimen collection/test date, or reported date.

### Outbreak-associated cases

Outbreak associated cases ≥14 years old were included based on work eligibility in Ontario.^14^ Demographic information included gender, age, presence of one or more comorbidities (list previously described)^9^, and Ontario region (Central East, Central West, Eastern, North East, North West, South West, Toronto). Clinical symptoms were classified as asymptomatic, presymptomatic (defined as having a testing date prior to symptom onset date), symptomatic, or missing. Clinical outcomes were classified as not hospitalized, hospitalized, intensive care unit without a ventilator, required a ventilator, and death. Outbreak cases were compared to sporadic (case not associated with any type of outbreak) cases ≥ 14 years old over the same time period in Ontario.

### Household transmission

We defined household spread using a natural language processing algorithm to link confirmed workplace outbreak-associated cases to other confirmed COVID-19 cases in Ontario by matching their residential address. Symptom onset dates of household and outbreak cases were compared to describe overall household transmission as: acquisition (household case was 2-28 days prior to outbreak case), transmission (household case 2-28 days after outbreak case), or unknown direction (household case ±1 day of outbreak case). Onset of illness was defined as symptom onset date, which was available for 60% of outbreak cases and 63% household cases. For those missing symptom onset date we calculated the weekly median number of days from test date to symptom onset date where data were available and performed a deterministic imputation based on the week of testing. If test date was not available we used the weekly median time from symptom onset to date reported to the local PHU to impute symptom onset. Onset date in asymptomatic individuals was the testing date or reporting date if testing date was not available. For any households with two or more cases associated with the same workplace outbreak, subsequent cases were considered due to household transmission to assess burden of illness from household spread.

### Statistical Analysis

Demographic, risk factor and outcome variables were compared between outbreak cases and sporadic cases by chi square test or Mann-Whitney test for non-parametric measures. Two sided p<0.05 was considered significant. All descriptive and statistical analyses were conducted in SAS version 9.3(SAS Institute, Cary, NC).

This analysis was approved by the Public Health Ontario Research Ethics Board (2028-020.01).

## Results

There were 199 workplace outbreaks declared by PHUs in Ontario during the study period, with 1245 outbreak-associated cases (**Table 1**). There were three or more outbreaks associated with nine industry sectors, with the majority in Manufacturing (45%), Agriculture, Forestry, Fishing and Hunting (12%), and Transportation and Warehousing (11%), with cases in these sectors accounting for 56%, 16% and 8%, of all outbreak cases, respectively. Outbreaks ranged in size from 1-140 cases, with 149 (75%) having two or more cases. Median outbreak size was largest in the Public Administration sector (9 cases) and smallest in the Retail sector (1 case), and while Manufacturing had the largest outbreak (140 cases), its median size was 3 cases. Outbreak duration ranged from zero days (50 outbreaks with a single case) to 119 days, with Agriculture, Forestry, Fishing, Hunting having the longest median duration (13.5 days), and the Retail sector having the shortest median duration (0 days), due to eight outbreaks with only a single associated case. Workplace outbreaks were infrequent from the end of February until mid-April 2020, and steadily increased from the beginning of May, when public health restrictions began to ease, to a peak of 8 outbreaks declared in a single day on May 27 (**Figure 1**).^1^ In comparison, sporadic (non-outbreak associated) SARS-CoV-2 cases in Ontario peaked in March, and then had a smaller peak in May (**Figure 1**). Both sporadic cases and workplace outbreaks steadily declined over June. There were no specific patterns of sectors affected over the time period.

**Table 1.** The number, size and duration associated with workplace outbreaks by industry sector in Ontario from January 21 to June 30, 2020, with associated cases until July 28, 2020.

| Sector Code | Sector | Outbreaks, n | Cases, n | Median Size (range) | Median Days Duration (range) |
| --- | --- | --- | --- | --- | --- |
| 11 | Agriculture, forestry, fishing and hunting | 24 | 197 | 3.5 (1-34) | 13.5 (0-119) |
| 23 | Construction | 12 | 44 | 2.5 (1-8) | 4.5 (0-12) |
| 31-33 | Manufacturing | 89 | 702 | 3 (1-140) | 9 (0-119) |
| 44-45 | Retail | 13 | 39 | 1 (1-14) | 0 (0-56) |
| 48-49 | Transportation and Warehousing | 22 | 103 | 2 (1-36) | 5.5 (0-75) |
| 52 | Finance and Insurance | 5 | 14 | 2 (1-5) | 6 (0-31) |
| 62 | Health care and social assistance | 13 | 43 | 3 (1-13) | 5 (0-50) |
| 72 | Accommodation and food services | 5 | 15 | 2 (2-5) | 6 (2-16) |
| 92 | Public Administration | 3 | 21 | 9 (2-10) | 10 (3-18) |
|  | Other Sectors with <3 outbreaks | 13 | 67 | 3 (1-16) | 7 (0-52) |
|  | **Total** | **199** | **1245** | **3 (1-140)** | **7 (0-119)** |

**Figure 1.**
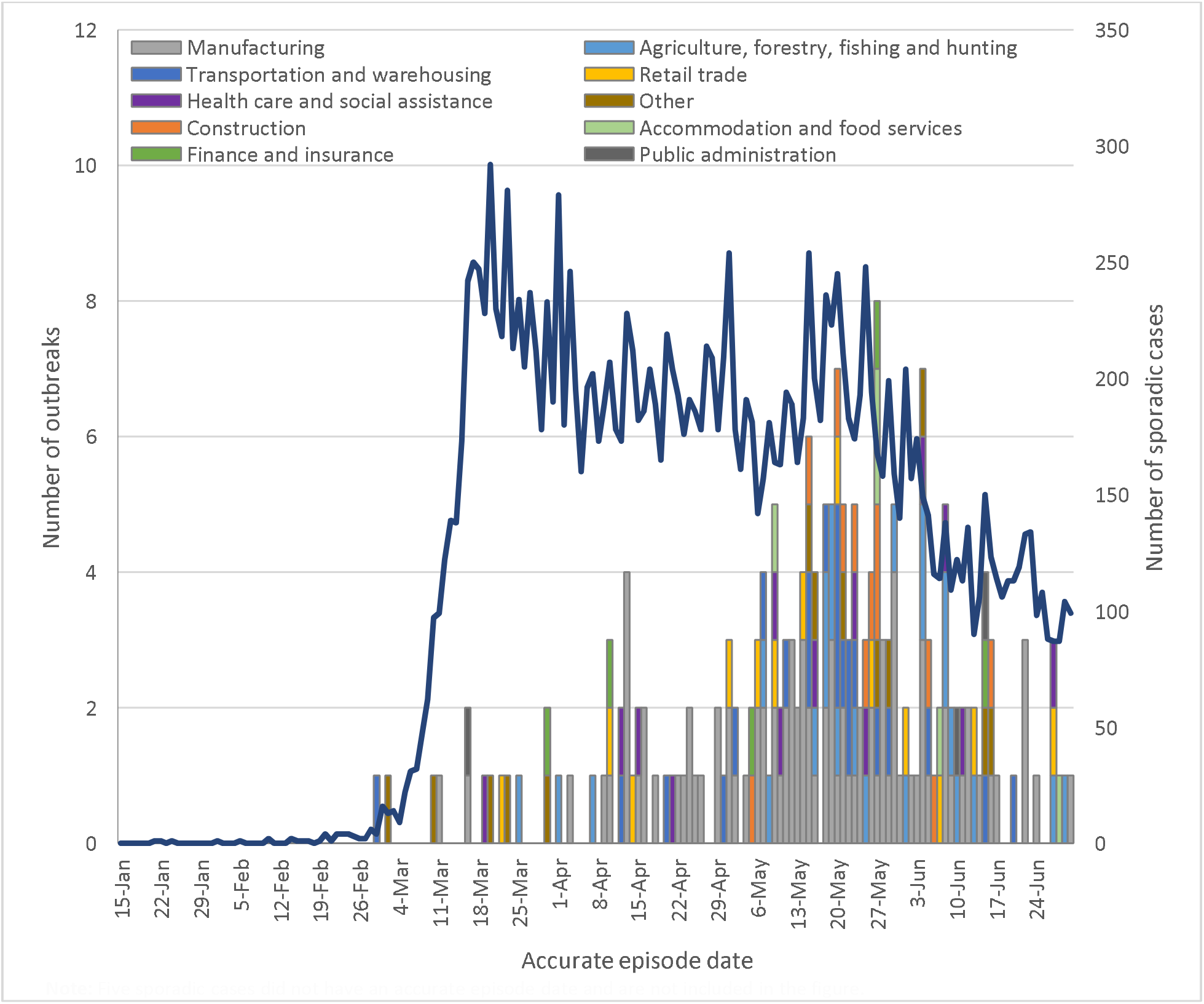
Distribution of workplace outbreaks by industry sector from January 21 to June 30, 2020, and the daily count of sporadic (non-outbreak) cases in that time period, Ontario. Note: Five sporadic cases did not have an accurate episode date and are not included in the figure

Outbreak associated cases compared to sporadic cases were significantly more likely to be male (71.7% vs. 51.5%), younger (median age 40 years vs 45 years), and were less likely to have one or more comorbidities (22.8% vs 33.1%) (**Table 2**). Outbreak cases were significantly more likely to be asymptomatic (20.7% vs 9.2%) or presymptomatic (3.9% vs 2.5%), and less likely to be hospitalized (4.2% vs 12.3%). There were a total of 52 hospitalizations and 6 deaths among workplace outbreak associated cases. Workplace outbreak cases were under-represented compared to sporadic cases in most regions of Ontario, but over-represented in the South West (17.4% vs 8.0%) region.

**Table 2.**
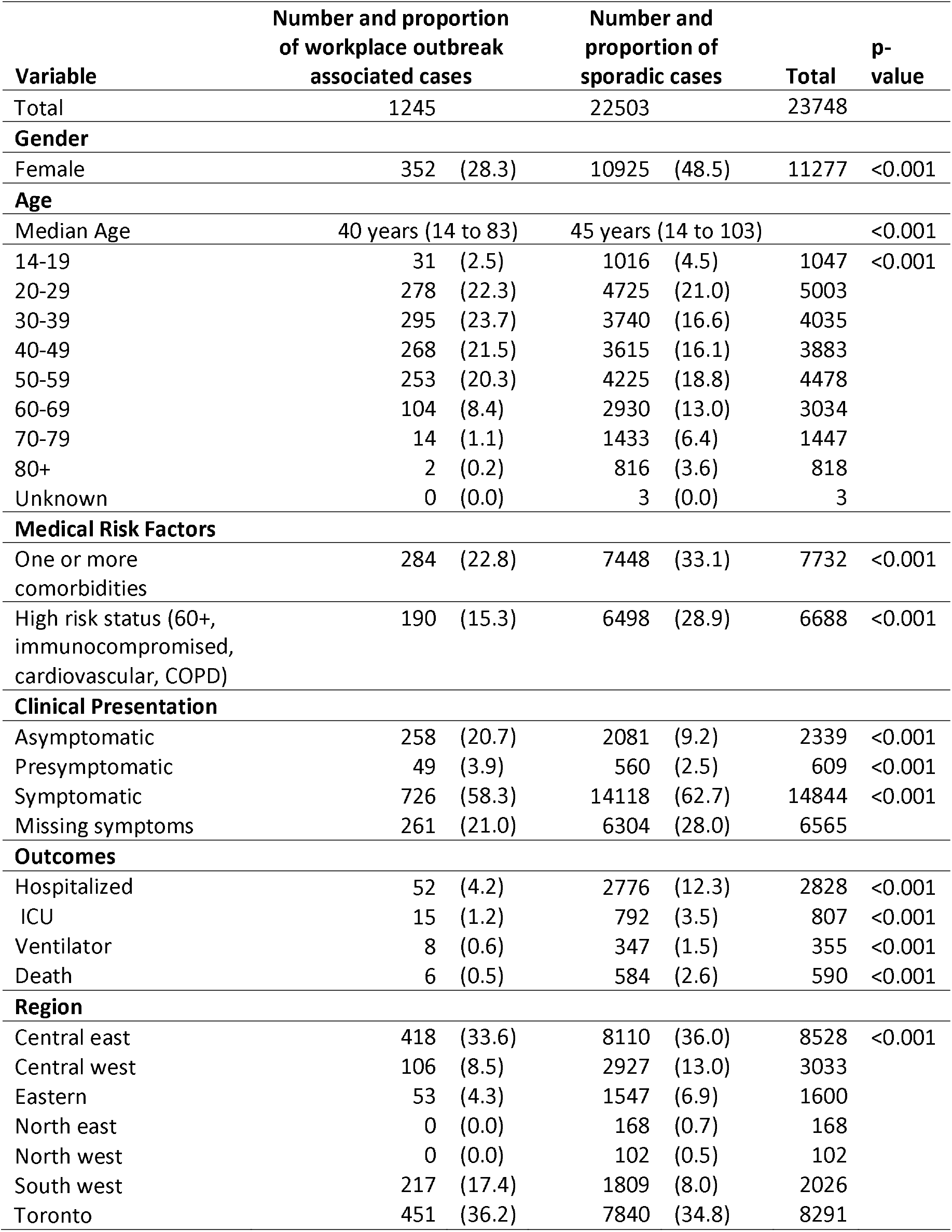
Demographics, medical risk factors, clinical presentation, outcomes and region of workplace outbreak associated cases and sporadic cases (non-outbreak associated) 14 years of age and older, Ontario, January 21 – July 28, 2020.

Among 1196 (96%) workplace outbreak cases with a valid residential address, 339 were matched to 608 household cases. There was a median of one household case associated with the 339 workplace outbreak case (range 1-12 cases). Of the household cases, 17% (103/608) belonged to the same workplace outbreak, but considered household cases as they occurred after the initial workplace outbreak case in their household, and leaving 31% (339/(1196-103)) workplace cases with household transmission. Therefore, accounting for household cases among cases with a valid address increased the burden of illness associated with workplace outbreaks by 56% (608/(1196-103)). Whereas assuming all cases are workplace associated (even if in the same household), the household transmission burden of illness increases by 42% ((608-103)/(1196)). Based on onset dates, 11% (66/608) of household cases were ≥2 days prior (acquisition), 60% (368/608) were ≥2 days after (transmission), and 29% (174/608) were ±1 day (unknown direction) relative to the outbreak case. Household cases were also most likely to be working age adults (19-59 years old) (74%), followed by <19 year olds (15%), and ≥60 year olds (11%) (**Table 3**). Female outbreak cases were associated with 33% of household cases, but only account for 28.3% of workplace outbreak associated cases. The proportion of outbreak cases with a corresponding household case ranged by industry sector, from 7% (Construction) to 40% (Accommodation and Food Services) (**Table 4**). Overall, the ratio of household cases to outbreak cases was 0.5, with the highest ratio by far in the Accommodation and Food Services sector (1.1), and lowest in Construction (0.1). The proportion of female cases and median age of cases was similar between outbreak cases with household cases and outbreak cases overall by industry sector, with no apparent trends across sectors between demographics of outbreak cases.

**Table 3.** Addressed matched household cases (n=608) by age group and timing of onset relative to their workplace outbreak associated case.

| Timing relative to associated workplace outbreak case | < 19 years | 19-59 years | ≥60 years | Total (%) |
| --- | --- | --- | --- | --- |
| ≥2 days prior (Acquisition) | 7 | 48 | 11 | 66 (10.9) |
| ≥2 days after (Transmission) | 60 | 261 | 47 | 368 (60.5) |
| ±1 day (Unknown) | 22 | 141 | 11 | 174 (28.6) |
| Total | 89 | 450 | 69 | 608 (100) |

**Table 4.** Description of workplace outbreak associated cases associated with an address matched household case(s) by industry sector.

| Sector code and name | Outbreak cases with household case relative to overall outbreak cases | % Female of outbreak cases with household case | Median age (range) of outbreak cases with household case | Household cases | Proportion of household cases to overall outbreak cases |
| --- | --- | --- | --- | --- | --- |
| **11. Agriculture, forestry, fishing and hunting** | 23/197 | 35% | 35 (16-62) | 40 | 0.2 |
| **23. Construction** | 3/44 | 0% | 43 (41-52) | 6 | 0.1 |
| **31-33. Manufacturing** | 227/702 | 26% | 45 (16-76) | 395 | 0.6 |
| **44-45. Retail trade** | 11/39 | 18% | 46 (28-74) | 24 | 0.6 |
| **48-49. Transportation and warehousing** | 20/103 | 45% | 40.5 (18-64) | 50 | 0.5 |
| **52. Finance and insurance** | 5/14 | 80% | 41 (26-55) | 7 | 0.5 |
| **62. Health care and social assistance** | 14/43 | 86% | 45 (18-83) | 23 | 0.5 |
| **72. Accommodation and food services** | 6/15 | 83% | 43.5 (21-53) | 16 | 1.1 |
| **92. Public administration** | 5/21 | 80% | 30 (28-65) | 7 | 0.3 |
| **Other sectors** | 25/67 | 32% | 51 (22-69) | 40 | 0.6 |
| **Total** | 339/1245 | 33% | 44 (16-83) | 608 | 0.5 |

## Interpretation

Three sectors (Manufacturing, Agriculture, Forestry, Fishing, and Hunting, and Transportation and Warehousing) accounted for two-thirds of the public health unit declared non-hospital, non-congregate, non-childcare workplace outbreaks of COVID-19 between January and June 30, 2020, in Ontario. While most outbreaks only had a few cases, the largest outbreak had 140 cases accounting for 11% of all workplace outbreak cases in this analysis. Accounting for household transmission associated with workplace outbreak cases with valid addresses increased the case burden of illness by 56%.

Analysis of workplace outbreaks by NAICS sectors in the United States found Manufacturing was also the most common sector for workplace outbreaks; however, Construction and Wholesale Trade were the next most common sectors in that study.^5^ Manufacturing, and particularly food manufacturing, is a sector that requires on-site work at specific times, and often in crowded conditions where physical distancing in lines of work is not possible, and environmental requirements for refrigeration/humidity may impact the use of personal protective equipment for COVID-19. Recommended prevention measures, such as plexiglass barriers where distancing is not possible, evolved over time as more was known about the risk of transmission in these settings.^15-17^

Farm-related outbreaks, as part of the Agriculture, Forestry, Fishing, Hunting sector, may be over-represented in Ontario, and in particular the South West region of Ontario, due to intense arable farming in the region.^18-20^ There was more than double the percentage of workplace outbreak cases in South West compared to sporadic cases in the region, largely due to the burden of farm-related outbreaks there, whereas most other regions had a lower proportion of outbreak cases to sporadic cases. As this analysis removed farms with evidence of congregate living (>4 cases with the same address), where there was significant spread among migrant farm workers living in dormitory-style housing, this analysis is an under-representation of the full burden of illness among farms in Ontario. Some farms with congregate living may still be included in this analysis. Outbreaks in this sector had the longest median duration, almost double than the overall median duration (13.5 vs 7 days), suggesting there may be different challenges in bringing outbreaks under control in this sector, and an increased impacts on workplaces and workers from extended application of outbreak measures. Farm-based mass testing programs may have also contributed to enhanced identification of farm outbreaks, as well as identification of asymptomatic cases. Overall, for all sectors, provincial workplace outbreak guidance recommending broad testing in the workplace once an outbreak is identified likely contributed to the increased proportion of asymptomatic and presymptomatic cases compared to sporadic cases.^12,21^

Workplaces in the Transportation and Warehousing sector often included both activities (trucking and warehousing). Warehouse conditions may face similar indoor crowded conditions as the Manufacturing sector. However, concern has also been raised by Canadian long-haul truck drivers that make frequent crossings into affected areas in the United States.^22^ Only select details on occupation are routinely collected for surveillance, and job type data were not available to assess relative contribution from Transportation vs Warehousing activities.

The timing of the peak of workplace outbreaks at the end of May is potentially linked to re-opening measures in Ontario starting from the beginning of May, thereby increasing the potential locations for workplace outbreaks to occur.^1^ However, updated data entry guidance for PHU reporting of workplace outbreaks was issued on May 8, potentially increasing the capture of outbreaks from that time (Public Health Ontario, 2020). Of note, the downward trend of workplace outbreaks paralleled declines in sporadic cases through June, and continued after issuance of provincial outbreak management guidance on June 11.^12^ Additionally, most education settings were virtual-only in the analysis time frame.

Workplace outbreak cases were younger, healthier, more likely to be male and had less severe outcomes compared to sporadic cases. Although there is evidence of a potential “healthy worker effect”, there were 52 hospitalizations and 6 deaths among included cases. Additionally, every case poses a risk of further spread in their home and community. While full contact lists for outbreak cases were not available to assess the overall community burden, our household transmission analysis suggests that initial cases in the household may be seeds for worker acquisition, introduction to the workplace, and subsequent transmission to other households. Therefore, household spread increases the case burden of illness related to workplace outbreaks, but also may increase the severity of impact if household members are more likely to be hospitalized or die from COVID-19 than the workplace outbreak cases. The proportion of workplace associated cases associated with household transmission (31%) was similar to what was found in another study among all cases in Ontario and other studies internationally.^23-26^ While there were no specific trends across sectors by age and gender for household transmission, further investigation could assess the apparent increase in household transmission among workers in the Accommodation and Food Services sector.

Limitations of this analysis include incomplete case capture where there were strict testing criteria early in the pandemic limiting detection of all cases, and broad asymptomatic testing and testing of close contacts was not introduced until the beginning of June, 2020.^1^ Additional cases may be missing linkage to an outbreak, under-representing the magnitude of workplace outbreaks. Manual assignment of outbreak locations may have led to misclassification by sector; however, this is less likely given broad sector categories versus analysis by subsector. Industry sector may not be representative of the actual occupation of outbreak cases. Use of address matching may have under-represented household transmission cases if addresses were missing, incomplete, or contained errors. Finally, estimates of workers by sector over this time period was not available for this analysis to assess case rates by sector.

## Conclusions

Workplace outbreaks have significant impacts on the workers and industries affected, as well as broader community impacts from household transmission. Sector-specific guidance is needed to address sectors at increased risk of outbreaks. As further loosening/tightening of public health measures continue over time, future analyses are needed to reassess trends in affected sectors when a greater number of sectors are operating, affected workers, and broader community transmission related to workplace outbreaks.

## Data Availability

Public Health Ontario (PHO) cannot disclose the underlying data. Doing so would compromise individual privacy contrary to PHOs ethical and legal obligations. Restricted access to the data may be available under conditions prescribed by the Ontario Personal Health Information Protection Act, 2004, the Ontario Freedom of Information and Protection of Privacy Act, the Tri-Council Policy Statement: Ethical Conduct for Research Involving Humans (TCPS 2 (2018)), and PHO privacy and ethics policies. Data are available for researchers who meet PHOs criteria for access to confidential data. Information about PHOs data access request process is available on-line at https://www.publichealthontario.ca/en/data-and-analysis/using-data/data-requests.

## Acknowledgements

The authors would like to thank Karen Johnson, Brenda Lee and Michael Whelan for their support in preparing the datasets for this analysis.

